# Walking the TB journey: Aboriginal lived experience shaping TB care in Northern Australia

**DOI:** 10.64898/2026.10.08.26365024

**Authors:** Emma L Smith, Chris Lowbridge, Jobe Bonney, Nikkita Joshua, Will Martin, Antonella Martin, Rachel Brian, Sonia Roberts, Charlie Gunabarra, Juan Darwin, Valda Bokmakarray, Kylena Rostron-Smith, Patricia Gibson, Jessica Gatti, Trang Nguyen, Vicki Krause, Elizabeth Stephenson, Jane Davies, Leisa McCarthy, Anna Ralph, Sarah Bernays

**Affiliations:** Menzies School of Health Research, Darwin, Australia; Centre for Disease Control, NT Health, Darwin, Australia; Sunrise Health Services, Katherine, Australia; Mala’la Health Service Aboriginal Corporation, Darwin, Australia; University of Sydney, Sydney, Australia

## Abstract

**Introduction:** Tuberculosis (TB) care is typically organised around biomedical and public health priorities, including diagnosis, infection control, treatment adherence, and prevention of transmission. Although qualitative research has described the burden of TB, lived experience is often treated as descriptive rather than as knowledge that can inform care. We examined how Aboriginal people in remote northern Australia experienced TB, and what these experiences reveal about how TB care can be strengthened in practice.

**Methods:** We conducted an inductive qualitative study involving in-depth interviews with Aboriginal people with lived experience of TB. Interviews were supported by Aboriginal community health workers. Data were analysed using reflexive thematic analysis, with Aboriginal contributors involved in interpretive workshops to refine themes and guide culturally safe representation.

**Results:** Twenty-one Aboriginal people contributed across 13 interviews. Three interrelated themes described the TB journey. Illness often challenged dominant biomedical expectations: symptoms were commonly normalised, which fear, ambivalence, and endurance were shaped by experiences of compromised health and cumulative loss. Distance from family, community, and Country was experienced as both protection and sacrifice; separation was endured to protect others but carried substantial costs, particularly when its purpose and duration were unclear. Recovery involved rebuilding strength, connection, and identity. Clear explanation, continuity, and opportunities for choice supported engagement in care, while survivors and community health workers provided important sources of trust, support, and shared responsibility.

**Conclusion:** Lived experience is practice-relevant knowledge for TB programmes. Care should minimise avoidable harms of separation, support continuity and choice, and recognise survivors, Aboriginal health workers, and communities as partners in the design and delivery of care.

## Introduction

Tuberculosis (TB) continues to be experienced in Australia, with Aboriginal communities bearing a disproportionate burden of disease and its consequences.[1, 2] TB disrupts everyday life, relationships, and connection to place, often requiring prolonged engagement with health systems and periods of mandated separation from family, community, Country and culture.[3]

Globally, TB care is typically organised through biomedical and public health models that prioritise diagnosis, infection control, treatment adherence for cure, and prevention of transmission. While these approaches are clinically essential, they can sit uneasily alongside the social and relational worlds in which illness is experienced.

A substantial body of qualitative research has documented the lived experience of TB globally, frequently describing stigma, fear, treatment burden, and social disruption.[4–6] While this work has been critical in humanising TB and highlighting patient perspectives, lived experience is often treated as illustrative rather than instructive, with limited influence on how care is organised, delivered, or evaluated in clinical practice.

In this paper, the term *relational* refers to understandings of health and care as embedded within relationships between people, and among family, community, and Country. Relational care refers to clinical practices that recognise and work within these relationships, including attention to family and community responsibilities, clear communication, and opportunities for choice and connection during care.

We explore how Aboriginal people experience TB across diagnosis, treatment and recovery, examining how lived experience can inform the design and delivery of TB care.

## Methods

### Study design and setting

This qualitative study explored Aboriginal people’s lived experiences of TB and how these experiences can inform the delivery of TB care in remote northern Australia. Interviews were conducted between October 2024 and August 2025 across multiple remote Aboriginal communities in northern Australia. The study formed part of a broader program of work focused on improving the quality of TB care and was conducted within a context of ongoing clinical and research engagement with participating communities, with relationships developed over time rather than established solely for the purposes of this study.

Participating communities ranged in population from approximately 100 to over 3000 residents and were located 400–650 km (5–8 hours’ drive) from the nearest metropolitan TB service hub. Access is predominantly by road and is seasonally restricted during the wet season. Travel between communities is common for family, cultural and healthcare reasons. While movement is a normal and valued part of social and family life, it can be logistically complex and, in the context of TB care, may become disruptive.

### People involved and sampling

Throughout this paper, the term *Aboriginal* is respectfully used to reflect the identities of the people and communities who shared their knowledge and experience. We use the term *people* in lieu of the more conventional ‘participants’, reflecting a relational approach to research and the ways individuals contributed as knowledge holders rather than solely as research subjects.

The research team included an infectious diseases physician and PhD candidate, a senior research fellow, and a remote area nurse, all with experience in remote TB care. An Aboriginal and Torres Strait Islander research assistant from the NT contributed to study implementation, selected interviews, analytic discussions, and theme development. Aboriginal community health workers and community contributors contributed to project design, interview development, recruitment, selected interviews, and interpretive workshops, and advised on cultural safety and knowledge that should remain within community rather than be published.

People were eligible if they identified as members of participating communities, could provide consent or assent, and had lived experience of TB disease within the previous three years, either as a person with TB or as a parent, guardian, or family member affected by TB. Recruitment was preceded by a period of community sensitisation and relationship-building undertaken in partnership with community health workers. This included community discussions about the purpose of the study, its relevance to TB care, and how findings would be used, to support transparency and trust.

Purposive sampling was conducted via regional TB clinic case and contact lists and community networks, seeking diversity in age, gender and experience of TB. People were approached by a member of the research team together with a local Aboriginal community health worker. Contact was relational and flexible, with time allowed for discussion and repeat visits before consent. To protect confidentiality in small remote communities, only broad descriptors are reported and communities are not named.

### Data collection

In-depth semi-structured interviews were conducted primarily by the lead author, with additional interviews by the senior research fellow and remote area nurse, alongside Aboriginal community health workers. Interviews used a conversational approach to support storytelling and person-led accounts (interview guide in Appendix 1). Conversations took place at times and locations chosen by the person, including at home, at the beach, or outdoors under a tree, and in preferred languages with local Aboriginal community health workers supporting interpretation where required. Interpreters were often known to the person and sometimes family members; interviews could become collective conversations reflecting local communication practices.

Interviews were audio-recorded. When interviews occurred in an Aboriginal first language, interpretation into English occurred during the interview, with transcripts produced from the English interpretation. Interviews were flexible in duration and sometimes conducted over more than one visit. Gender matching of interviewer was offered and was taken up in several cases; in others, people preferred a same-gender community member to be present. To minimise power imbalances, researchers did not interview anyone for whom they had current or prior involvement in clinical management. People were offered a token of appreciation (AUD $40 store voucher or phone credit).

### Data management and analysis

Data were analysed using reflexive thematic analysis with an inductive orientation, with coding and theme development grounded in people’s accounts rather than a pre-existing theoretical framework. Analysis was iterative, involving repeated engagement with the dataset and review of developing themes against coded extracts and the full dataset. Aboriginal community health workers participated in interpretive workshops to develop, refine, and contextualise themes and to identify knowledge shared in trust that was not considered appropriate for publication or broader dissemination.

The lead author is a non-Indigenous infectious diseases physician working in remote TB care. This positioning was recognised as shaping relationships with people involved in the study and interpretation of the data, and reflexive attention was maintained throughout analysis, including through iterative discussion with co-authors and Aboriginal community contributors (full reflexivity statement in Appendix 2).

### Ethical considerations

Approvals were obtained from Human Research Ethics Committee of NT Health and Menzies School of Health Research (Ref: 2023-4752), and the boards of participating Aboriginal Community Controlled Health Services.

## Results

Twenty-one Aboriginal people took part across 13 interviews. Ages ranged from 19 to 68 years and included twelve women and nine men. Twelve had lived experience of TB disease and nine were family members involved in care and support. Among those with lived experience, some were interviewed during treatment, while others had completed treatment within the previous three years. All lived in remote Aboriginal communities in northern Australia.

Three interrelated themes were developed to describe how TB was experienced and interpreted, how separation and distance were navigated, and how recovery and return were understood. While presented separately for analytic clarity, the themes are closely interwoven. Together, they show how lived experience exposes tensions within dominant models of TB care and points toward more responsive practices.

### When illness expectations collide with TB care

This theme describes how living with TB unfolded within everyday experiences of illness, uncertainty, and loss that often differed from assumptions embedded within clinical and institutional models of care. Expectations about symptom seriousness, urgency, prognosis, and recovery were not always shared between people experiencing TB and the health systems providing care. People spoke of tolerating symptoms and continuing daily life while unwell, with seriousness not always immediately apparent. Illness was encountered in a context where serious illness and death were familiar rather than exceptional, shaping fear, ambivalence, and endurance in ways that differed from expectations of early recognition and straightforward recovery. These differing expectations created tensions with healthcare systems that rely on urgency, and recovery as the expected trajectory.

#### Illness that is lived with and normalised

Early TB symptoms were frequently normalised, particularly when daily function could still be maintained. Ongoing discomfort and reduced health were described as common experiences, meaning symptoms were often interpreted as manageable rather than serious. This was described by one midlife female family member: *‘What I noticed is that my son had a back pain, always, he had back pain. That’s why I didn’t realise that he had that TB. I thought that it was a normal back pain.’*

TB was experienced less as the presence of ‘typical’ symptoms and more as the gradual loss of everyday functioning and independence. Although illness developed gradually, people often adapted to worsening symptoms until everyday functioning could no longer be maintained, making the transition from manageable to unacceptable feel sudden.

> ‘*I was weak and couldn’t do anything. Couldn’t get up. Or actually go to the toilet or shower or anything. I was just laying down.’* (Young female, lived experience B)

Some people later reflected that although they recognised the diagnosis when it was explained, they were not emotionally ready to accept it at the time.

> *‘I was like…”No that can’t be. It’s not real.” […] I was telling them; “this is a lie”. But I knew it was real. I was just angry and anxious.’* (Young male, lived experience B)

This reflects denial as a protective response as distressing information is gradually processed, particularly when symptoms had previously been understood as “normal.”

When experiencing symptoms, people often responded first within local and familiar systems of care, including family support, traditional healing practices, and remote community clinics. Early presentations were often interpreted as more common illnesses, reflecting the non-specific nature of TB symptoms. In several accounts, seriousness was only felt to be recognised once referral outside the community was arranged, with transfer to hospital understood as confirmation that something more serious was occurring.

#### Death, grief & expectations of seriousness

TB was interpreted within broader experiences of cumulative loss and frequent exposure to death. Serious illness was often assumed to lead to decline rather than recovery, shaping responses that ranged from intense fear to quiet acceptance.

> *‘I guess with our mob it takes time when we grieve. Especially losing loved ones after loved ones. It is hard. […] There’s been 3 or 4 deaths already this year.’* (Midlife female, family member F)

> *‘Every day I just lay down and was like, I’m not going to do anything about it. Just let it be and just pass on. A lot of young people my age die you know. That’s what I was thinking. About my life and that.’* (Young female, lived experience B).

Rather than indifference, this reflected an expectation, grounded in lived experience, that serious illness often led to death and outcomes were not easily altered. The diagnosis was therefore interpreted within a framework where mortality was plausible and recovery not assumed.

For some people, the death of a family member from TB fundamentally changed how the illness was understood. TB was no longer experienced as a distant or manageable illness but as one associated with profound loss, reshaping expectations about seriousness and survival.

> *‘It was very sad. Especially you know, sudden like that, and unexpected. I didn’t think TB was that dangerous.*’ (Midlife female, family member F)

#### Deciding what to share and what to protect

In contexts where understandings of TB were inconsistent, people described carefully managing who was told about their diagnosis and why. Disclosure decisions were shaped by concerns about misunderstanding and relational responsibility, rather than automatic fear of stigma. Stigma and shame were not always explicitly named and were experienced unevenly. Several people suggested that TB carried less stigma precisely because it was poorly understood, contrasting it with conditions linked to moralised explanations, such as uncleanliness, which were more clearly associated with shame.

> *‘Most people don’t know anything about TB, so they wouldn’t have any shame at all. I was living with a family with TB, and I didn’t even know what was that. And it wasn’t shame job’.* (Midlife female, family member D)

In this sense, shame and social discomfort appeared to emerge less from illness itself than from partial knowledge that positioned individuals as responsible for infection or its spread. Practices of selective disclosure and emotional privacy were not specific to TB alone, but reflected a broader preference to keep illness and personal circumstances discrete. These accounts also challenged assumptions that illness is always managed collectively within community, highlighting the ongoing importance of privacy, personal boundaries, and individual choice within relational worlds.

> *‘I didn’t want to do interview in front of them* [other members of the research team]*.[…] It’s just my feelings and my ways that I want things to be private’.* (Young male, lived experience B).

Within this context, some people also described deliberately normalising TB, framing it as one illness among many, as a way of resisting shame and reducing moral weight. Others chose openness within trusted relationships, where sharing was understood as a responsibility to protect others and enable support. Decisions about disclosure were often made collectively within families and Kinship networks.

> *‘We were spreading that news about it* [TB diagnosis]*. Telling all of the families’*. (Young male, family member)

#### Living with uncertainty: explanation, endurance, and relational care

Many people described ongoing uncertainty after diagnosis, including how TB was acquired, how long treatment would take, and whether recovery was likely. Where treatment purpose, duration, and expected outcomes were not clearly explained, uncertainty reinforced existing expectations that serious illness leads to decline rather than recovery.

> *‘I want to know how you get the TB. We don’t even know how. We have requested that*.’ (Midlife female, family member E).

Some people expressed concern about medication or a desire to return to bush medicine, particularly early in treatment. Medication side effects were commonly experienced, often downplayed relative to everyday bodily discomfort, and did not automatically prompt treatment interruption. However, continuation was not assumed; willingness to persist depended on whether treatment was understood as restorative and time-limited. Where this was clearly explained, side effects were more readily endured.

> *‘I didn’t vomit or feel sick inside my stomach. I was holding in my hand, like 20 tablets.’* (Young female, lived experience A).

Treatment was often accepted gradually over time, as people worked through fear, uncertainty, and the implications of diagnosis. These accounts suggest that beginning treatment was not always an immediate decision, but a personal process of reflection, adjustment, and growing acceptance.

> *‘I thought I wouldn’t end up taking the medication, but it was hard for me. When I came back home, I had to always think about it. How to accept the medication. I had to think about it for days. Then I called and said I was so sorry and to let them know I will take the medication. That’s when I started’. (*Young male, lived experience B).

Across accounts, harm associated with TB care was reduced when care made space for explanation, time, and relational presence. Many people began treatment from a position of fear and limited expectation of recovery. In this context, consequence-focused or fear-based messaging often reinforced doubt rather than motivating action. Explanations that emphasised restoration, recovery, and purpose were more reassuring and better supported engagement.

Being given time to process information, rather than being expected to understand or decide immediately, was consistently described as respectful and supportive.

> *‘We go twice then come back, so they can think […]. Giving people time to process. We just sit down and have a yarn. Sit down and talk to them. Talk to those sick people*.’ (Midlife female, lived experience A)

People also emphasised the importance of healthcare providers consistent presence in community, describing how trust and reassurance were built through continuity and visibility rather than one-off encounters. Ongoing presence helped counter perceptions that care was disconnected from local relationships, creating a sense of reciprocity in a system otherwise experienced as one-sided.

> *‘Come visit every month. It would be good for TB mob to come visit every month. Spending more time in the community would be great.’* (Mid-life female, family member D)

### Distance as protection and sacrifice

This theme describes how TB required physical separation from family, community, and Country, experienced as both protection for others and profound loss. Distance was often endured to protect others, reflecting collective responsibility and sacrifice. At the same time, separation carried significant emotional, relational, and existential costs, including loss of freedom, privacy, and sense of self. These accounts show how being kept apart was lived as a necessary form of care that could not be disentangled from harm, revealing the moral weight distance carries.

#### Separation as protection and loss

Separation from family, community, and Country was described as distressing, bringing feelings of fear and loss that were difficult to disentangle. While travel outside community was common for many people, enforced stays for hospital-based treatment, often of uncertain duration, were experienced differently. Being removed to distant hospital settings created vulnerability and uncertainty.

> ‘*It was hard. It is actually my first time. Going away for 3 weeks. Staying in the hospital. […]. Really hard…devastating.*’ (Young female, lived experience B)

People also described actively managing physical and social closeness, with separation sometimes chosen or supported by families to protect others. In this way, distance was understood not simply as loss, but also as protection.

> *‘Well, the thing is you can’t share things to the person that has TB. Or even when the person is sitting next to you, they shouldn’t cough towards you, because their germs come to you. Maybe they don’t want to come back and stay with family and spread it. That way they can be there alone to be safe. And not spreading to anyone.*’ (Midlife female, family member C)

Remaining in hospital was often described as a difficult but necessary sacrifice. The value of treatment was frequently understood in relational terms, as protecting family and community from harm, rather than primarily as supporting their own recovery.

Hospitalisation was associated with reduced freedom and control, with people describing feeling managed and directed by healthcare systems. A midlife woman with lived experience described her frustration at mandated isolation in hospital*: ‘It was hard to get out. Because of the doctors and nurse. They wouldn’t let me get out. But I didn’t want to be there’*.

Further accounts reflected a sense of obligation to do what healthcare staff advised:

> ‘*They told me, we had a look, and you’ve got that TB, and I was crying you know, and they wanted to keep me there, so I had to stay there*.’ (Older male, lived experience C)

While some welcomed protecting the community, separation was rarely experienced as beneficial to them personally. Isolation in hospital, reduced agency, and limited explanation were often described as burdensome and frightening. In this context, being required to endure separation and uncertainty eroded people’s sense of control over their care.

#### Choice and relational care reshape the experience of distance

Over time, for some people trust was rebuilt through care characterised by kindness, respect, and efforts to understand people’s concerns. These relational connections provided safety, reassurance, and confidence beyond information alone.

> *‘When she* [nurse] *came back, she would always come in and say, ‘I’m back’. And then she came in and gave her a hug in the bed. She does that every day when she does her shift.’* (Midlife female, family member E)

Accounts highlighted the importance of flexible care that supports choice and control. Choice strengthened dignity and agency, especially when loss and disruption were acknowledged. Having some choice in how and where treatment was delivered helped people feel recognised and supported rather than directed through care.

> *‘If I would be ending up with TB, I would rather choose community. And at least I would take the medicine, because I choose.’ (Midlife female, family member D)*

### Coming back strong – Rebuilding connection through recovery

This theme describes recovery as a process of regaining strength, agency, and connection over time. Treatment was often experienced as a turning point that enabled re-engagement with everyday life, supported by family, community and relational care. Recovery was understood as returning strong to Country and community, where healing, support and responsibility were shared.

#### Recovery as strength, identity and return

Over time, many people came to see the disruption and sacrifice of treatment as worthwhile, as their physical health began to improve. People described feeling stronger, or more like themselves after starting TB medication. For many, this visible improvement marked a clear shift in how treatment was understood.

> *‘What I’ve noticed is that his weight has started to go up. And also, that I’ve realised that he can walk straight and he’s getting strong and healthy by taking those medicines’.* (Midlife female, family member C)

Understanding why treatment was needed and what to expect helped people feel more in control within an otherwise demanding process.

> *‘I had to understand to take the medication and then I can go back to my family. So, I was understanding that’s the reason I was taking.’* (Midlife female, lived experience A)

People’s stories of recovery emphasised survival, renewed strength, and changed identity. As improvement became visible, confidence grew, often unexpectedly, given earlier assumptions that serious illness would lead to decline rather than recovery. Recovery became a source of pride, linked both to protecting family and community and to regaining personal wellbeing.

> *‘Yes, well to stay healthy you got to look after yourself. […] I didn’t want to let myself down. I just wanted to… take all my medicine, take all of the tablets and get back home. And I can enjoy my life again. And I did what I did’* (Older male, lived experience A)

Family and community support were central to endurance and recovery, with shared responsibility and collective pride frequently described.

> *‘I made it. It was because of the support I was getting. She* [mother] *was like, very supportive. And I don’t know what I would do without mum.’* (Young male, lived experience B)

> *‘They are both fighting well and I’ve seen them; they are strong and healthy right now. And I’m very happy for them.*’ (Young male, family member)

Recovery was described as a process of regaining health, routine, and a sense of self, rather than a sudden or complete resolution of illness.

> *‘I felt like me again. I tell them it’s the only way to make you better. To be the old you again*.’ (Young female, lived experience B)

Survivors reflected on their endurance through a difficult journey and a sense of change and renewed identity:

> *‘It really touched me when I got better you know. And now I don’t like seeing people suffering*. […] *After I got really sick, I started looking out for other people. Because I didn’t want them to go the same way I went.’* (Young female, lived experience B)

These accounts close with survival not only as recovery, but as a sense of responsibility to prevent others from suffering.

#### Recovery as collective care

People with lived experience of TB were seen as well positioned to support and encourage others. Although disclosure is not always routine, people described how sharing experience could provide reassurance and hope, challenging traditional hierarchies of expertise and highlighting the value of lived knowledge in community-based TB care.

> *‘They need that kind of experience, like leadership. So, he can tell them his side of the story. And tell them that he went through his own thing. And made it out. That’s how people can be brave. Coming and taking the right medicines*.’ (Young male, family member)

Community health workers were described as critical relational bridges between families, communities, and the health service. They are trusted figures who translate meaning, support understanding, and sustain continuity across care settings. A family member affected by TB who worked in this role expressed pride in contributing to community wellbeing.

> *‘Normally we can just take the medicine out there, to their homes. Have a little bit of a check with them. Talk about how they are going, how they are doing, and how to use the medications. That way we are very brave for them.*’ (Young male, family member)

The final accounts move to survivors’ own reflections on what supports recovery and what TB care should enable:

> ‘*We can do that by taking people out fishing, hunting, camping. You know…eating healthy food out bush. Keeping them safe. That’s the one. That always makes people healthy and doesn’t worry about anything. They eat bush foods or sometimes even bush medicine too. And then they come back and take balanda [*non-Indigenous*] way medicine too. That’s how we make them keep going strong and healthy*.’ (Young male, lived experience B)

> ‘*Because I had that medicine, I had bush tucker, family close to me. Everything*.’ (Midlife female, lived experience A)

## Discussion

Drawing on the experiences of Aboriginal people in remote Northern Australia, this study examined how people navigated TB diagnosis, treatment and recovery. Rather than treating lived experience as contextual background, this analysis positions it as evidence for organising and delivering TB care, reflecting broader shifts towards understanding healthcare as relational, context-dependent and complex.[7, 8] Across the findings, experiences of TB were shaped by prior expectations of illness, recovery, and care. Care was more readily experienced as supportive when explanations engaged with these understandings and positioned treatment as restorative, purposeful, and time-limited.

A central finding was that prior experiences of serious illness and cumulative loss shaped what a TB diagnosis and its treatment demands could signify. Fear and ambivalence occurred in a context where illness was often familiar and recovery could not be assumed. This pattern aligns with Aboriginal social and emotional wellbeing literature describing cumulative and collective grief, and with the concept of “bereavement overload” in global grief literature, where repeated loss can shape expectations of illness and survival.[9, 10]

A related but distinct finding was the normalisation of symptoms and discomfort within everyday experiences of health. Medication side effects were often endured rather than prompting treatment cessation, differing from many TB accounts in other settings.[6, 11] Similarly, early or non-specific TB symptoms could overlap with other prevalent conditions and be normalised or overlooked by community people and clinicians, a pattern also described for other endemic infectious diseases such as scabies.[12] Together, these findings suggest that familiar experiences of ill health may influence both when symptoms are recognised as requiring attention and what discomfort is considered tolerable during treatment. This cautions against interpretations of diagnostic delay or treatment engagement that locate the problem solely within individuals, rather than within the broader health contexts in which TB occurs.

Experiences of distance and separation further highlight the ethical complexity of TB care. Distance from family and Country was often endured as an act of protection and collective responsibility, but also as profound loss. Consistent with other TB literature, distancing was understood as protective of others,[13] yet here it was frequently experienced as a one-sided sacrifice because personal recovery or benefit from treatment was not always assumed. Where serious illness is commonly associated with decline or death, separation may be interpreted as open-ended unless its purpose, time limits, and personal health benefits are clearly explained. Relational care and opportunities for choice softened some of the harms of separation, but did not remove them. Being kept apart was experienced as a morally significant form of care requiring careful explanation of both collective and individual benefit.

TB stigma is highly context-dependent. In some settings it is strongly shaped by HIV association, while in others it attaches more closely to poverty or social marginalisation. Global qualitative evidence shows that TB stigma often follows existing axes of disadvantage.[14, 15] In this study stigma was not uniformly central to people’s experience. Decisions about disclosure were governed more by relational responsibility than by fear of judgement. This challenges assumptions that stigma is always the primary social barrier in TB care.

Across themes, explanation, time, relational engagement, and continuity were not simple valued features of supportive care; they shaped whether TB care made sense within people’s existing understandings of illness and recovery. These findings add context to person-centred TB care models emphasising communication, dignity, [16, 17] and evidence supporting continuity in remote health service delivery.[18, 19] When illness is understood through everyday experience, explanations based primarily on future risk may be carry limited meaning, particularly for preventive treatment of an invisible condition such as TB infection. In contrast, visible and continuous care, particularly when delivered on Country, strengthened trust and understanding. Improving TB outcomes therefore requires attention not only to delivering effective treatment, but also to how care is explained, experienced and supported (see Box 1).

Early clinical conversations should explore how diagnosis, treatment, and recovery are understood and revisit these understandings over time. Importantly, these findings do not imply that there is a single culturally appropriate model of TB care for Aboriginal people. People differed in the kinds of support, privacy, spirituality, family involvement, and treatment arrangements they valued. Culturally safe care therefore requires flexible, individualised approaches that create space to understand each person’s priorities, relationships, and explanatory frameworks. Clear explanation that treatment is time-limited and restorative for both individual and community can help recalibrate fear-based interpretations of treatment and separation.

Recovery was described not only as individual improvement but as relational and responsibility-bearing. Survivorship carried pride and obligation, with people identifying roles in supporting and educating others. In communities where serious illness was often associated with decline or death, visible experiences of recovery carried particular significance. Survivors provided tangible evidence that treatment could restore health and connection, helping reshape expectations of prognosis and possibility within families and communities. While survivor-led approaches are increasingly recognised in global TB advocacy, they remain underrepresented in TB literature.[20, 21] Peer-support approaches have demonstrated the value of lived experience in other infectious disease settings,including hepatitis B and C.[22, 23] Community-led models and Aboriginal health services similarly emphasise the importance of local leadership.[9, 24] In this study, community health workers were described as essential relational bridges between clinical and community worlds. Ending TB transmission and supporting recovery depends not only on clinical expertise, but on recgonising and resourcing survivorship and community-based leadership as central to care. [25]

This study was strengthened by sustained partnership with Aboriginal community members and health workers, who shaped recruitment, interview processes, and interpretation. The work was conducted in two regions of the Northern Territory of Australia, and the perspectives shared reflect those contexts rather than all Aboriginal communities.

While grounded in the experiences of these communities, the findings have relevance wherever TB care requires prolonged treatment, separation from family, or navigation between different understandings of illness and recovery. Lived experience offers more than an account of receiving care: it provide knowledge about how care is interpreted, what makes its demands meaningful or burdensome, and how recovery can become visible and credible to others. Culturally safe TB care therefore depends not only on what is delivered, but creating space for this knowledge to shape how care is explained, organised, and supported over time.

### Box 1: Practice implications for TB care

- Begin by exploring how the person understands their illness, its seriousness, and what recovery might mean to them
- Work toward shared understanding of illness, treatment, and recovery so that the clinician and patient are working from a shared understanding
- Avoid assuming a single culturally appropriate model of care; explore preferences for privacy, family involvement, spirituality, community-based care, and treatment support on an individual basis
- Frame treatment as restorative and time-limited, clearly explaining what treatment is likely to change, including potential improvement as well as side effects, for the individual, their family and the community
- Explain separation for care transparently, including its purpose, expected duration, and anticipated health benefit
- Move beyond one-off explanations with ongoing conversations that revisit and realign expectations over time and build trust
- Embed community health workers and people with lived experience within TB care to strengthen trust, continuity, and understanding

## Data Availability

The qualitative data generated in this study are not pubicly available due to the sensitive and personal nature of the interviews and the risk of identification within small remote communities.

## Acknowledgements

We extend our sincere thanks to the people who shared their experiences and knowledge with us. Their generosity and openness in sharing deeply personal stories were fundamental to this work. We are grateful for the trust placed in us to take these stories forward, and we recognise their contribution as vital to shaping care in Northern Australia and in other settings where these insights may resonate.

We also acknowledge the Aboriginal Community Health Workers who supported this research through their relationships, cultural guidance, and contributions to data collection, interpretation, and reflection. We are grateful to the Aboriginal Health Services and their Boards for their support of this work and for enabling it to take place in partnership with community.

We are especially grateful to Sophie Carey, Remote Area Tuberculosis Nurse, whose deep commitment to community, generosity, and support were central to this work. Her insight and guidance have shaped this research in countless ways, and this work would not have been possible without her.

